# Anti-Müllerian hormone is decreased in women with superficial peritoneal endometriosis and associated with an elevated inflammatory profile

**DOI:** 10.1101/2025.08.25.25334349

**Authors:** Meaghan J Griffiths, Martha E Brown, Douglas A Gibson, Frances Collins, Cheryl E Dunlop, Philippa TK Saunders, Andrew W Horne

**Affiliations:** EXPPECT Edinburgh, Centre for Reproductive Health, Institute for Regeneration and Repair, University of Edinburgh, UK; Edinburgh Fertility Centre, Royal Infirmary of Edinburgh, UK

**Keywords:** Endometriosis, infertility, AMH, ovarian reserve, superficial peritoneal endometriosis, inflammation, cytokines

## Abstract

**Study question:** How are the levels of anti-Müllerian hormone and inflammatory cytokines influenced by superficial peritoneal endometriosis (SPE)?

**Summary answer:** Fertility metrics (Endometriosis Fertility Index (EFI), and serum anti-Müllerian hormone (AMH) levels) are reduced in women with SPE. Simultaneously, inflammatory markers are elevated in the circulation and local pelvic peritoneal microenvironment, with distinct changes in each compartment.

**What is known already:** Between 25-40% of women with endometriosis experience infertility, though the mechanisms behind this are poorly understood. Ovarian endometriosis is known to decrease AMH levels and contribute to infertility, but little is known about SPE-associated infertility, and how the peritoneal microenvironment might play a role in infertility for women with SPE.

**Study design, size, duration:** Venous blood samples from women with suspected endometriosis were collected prior to diagnostic laparoscopy (n=105). Pelvic peritoneal fluid was also collected from a subset of the women (n=38). The Endometriosis Fertility Index (EFI) was calculated after surgery, and levels of AMH and inflammatory cytokines measured by ELISA or multiplex Luminex.

**Participants/materials, setting, methods:** Based on their surgical findings, women were classified as no endometriosis observed (no endo; n=39), superficial peritoneal lesions only (SPE; n=43), or SPE with an ovarian endometrioma (SPE+OE; n=23). Women were further grouped by their use of hormone treatments to manage their endometriosis symptoms (no endo: no hormones n=14, hormones n=25; SPE: no hormones n=20, hormones n=23; SPE+OE: no hormones n=17, hormones n=6). Data are described as either mean ± standard deviation, or median [interquartile range].

**Main results and the role of chance:** SPE+OE women were older (31.73±6.31) than SPE (27.77±6.14; p=0.04) and control women (27.65±5.81; p=0.02). Both SPE and SPE+OE groups had lower EFI scores compared to women with no endometriosis (no endo 9.41±0.50; SPE 8.63±1.11 p=0.04, SPE+OE 6.95±1.60 p<0.0001). Serum AMH levels were lower for SPE alone (p=0.009) and SPE+OE women (0.73ng/mL [0.32, 1.19], p=0.002) compared to women with no endometriosis (1.15ng/mL [0.75, 1.94]) when accounting for age. When also accounting for hormone use, women with SPE+OE had lower AMH levels compared to women with no endometriosis (p=0.02), while women with SPE alone did not (p=0.069). Moreover, women with SPE not using hormones had elevated serum IL-17 (4.45pg/mL [4.26, 4.88] vs 3.84pg/mL [3.54, 4.19], p=0.02) and TNF-α compared to women with no endometriosis (4.28pg/mL, [3.37, 5.88] vs 1.99pg/mL, [1.49, 3.43], p=0.03), while pelvic peritoneal fluid levels of IL-23 were elevated in women with SPE not using hormones (212.4pg/mL, [184.0, 244.5] vs 121.3, [46.37, 147.60], p=004). These differences were not significant in women using hormones.

**Limitations, reasons for caution:** Due to the limited sample size of women not using hormones, we were unable to determine if serum IL-17 or TNF-α, or pelvic peritoneal IL-23 levels negatively correlated with AMH levels.

**Wider implications of the findings:** Women with SPE, with or without OE, have lower AMH levels - indicative of reduced ovarian reserve - compared to women without endometriosis. Among those with SPE, diminished AMH was associated with increased serum levels of IL-17 and TNF-α and elevated IL-23 in the pelvic peritoneal fluid, suggesting compartment-specific inflammatory profiles. Notably, changes to circulating inflammatory cytokines were different when use of hormonal therapy was taken into account, highlighting such treatments may modulate inflammation linked to endometriosis. Taken together, our data support the need for further investigation into inflammation as a potential mechanism underlying infertility in women with SPE in the absence of OE.

**Study funding/competing interest(s):** Deanery of Clinical Sciences Funding Challenge, University of Edinburgh awarded to MJG.

**Trial registration number:** University of Edinburgh Lothian Ethics Committee REC 20/LO/1298.

## Introduction

Endometriosis is a chronic, hormone-dependent neuroinflammatory disease where endometrial-like tissue grows outside the uterus. It affects about 10% of reproductive age women worldwide, and currently, there is no cure (As-Sanie, Mackenzie et al. 2025). The four subtypes of endometriosis - superficial peritoneal (about 80% of cases), deep, ovarian (endometriomas), and extrapelvic - can occur alone or together. Superficial peritoneal endometriosis (SPE) is typically located on the surface of abdominal or pelvic organs and the pelvic wall. Deep endometriosis invades pelvic tissues or organs, like the bowel or bladder. Ovarian endometriomas (OE) are cysts lined by endometrial tissue within the ovary. Extrapelvic endometriosis involves lesions outside the pelvis and can affect many organs, including the diaphragm, thoracic organs, or even the brain.

A frequent symptom for up to half of women with endometriosis is infertility. Alone, infertility has detrimental impacts on mental health and quality of life for those wanting to conceive, which is further increased by endometriosis (Mori, Zaia et al. 2024). Clinically, both endometriosis and infertility suffer delays with diagnosis and a lack of effective, accessible, treatment options. In international guidelines for endometriosis, it is recommended women with SPE consider surgery to remove SPE lesions based on evidence that this may improve spontaneous pregnancy rates within the first 12 months post-surgery (NICE 2017, Becker, Bokor et al. 2022). In many UK centres, the waitlist for a diagnostic laparoscopy for endometriosis is two or more years (Endometriosis UK 2024, Royal College of Obstetricians and Gynaecologists 2024). For these women, assisted reproductive technologies (e.g. IVF) provide one potential solution, although this is also subject to access to NHS provision (in the UK), and is costly. A 2023 study found 3.2 million women of reproductive age in England had no or limited access to a fertility clinic in their area of residence, while the highest household income areas had the best access to fertility clinics (Jones, Peri-Rotem et al. 2023). Importantly, there is a group of women who are only diagnosed with endometriosis when they attend fertility clinics when seeking to become pregnant and the majority of these are cases of SPE (Van Gestel, Bafort et al. 2024). If these women are of advanced maternal age, it is unlikely that they will want to wait several years for a laparoscopy to remove the endometriosis in the hope it may enable them to conceive spontaneously. To provide better and more equitable options for these women, we must better our understanding of the mechanisms behind how SPE contributes to infertility.

The Endometriosis Fertility Index (EFI) predicts post-surgery pregnancy success for women with endometriosis based on the surgeon’s assessment of damage to Fallopian tubes, fimbria and ovaries, endometriosis staging score, patient age, and pregnancy history (Adamson and Pasta 2010). Since its inception, it has been validated in a variety of settings and shown to accurately predict non-IVF pregnancy success for women with endometriosis post-surgery (Tomassetti, Geysenbergh et al. 2013, Garavaglia, Pagliardini et al. 2015). It has also recently been shown it can be accurately completed without surgical intervention (Tomassetti, Bafort et al. 2021). However, to date, its utility has been limited to testing on combined datasets containing all subtypes of endometriosis.

In clinics offering assisted reproductive technologies, circulating levels of anti-Müllerian hormone (AMH) are typically measured to predict the capacity of the ovary to respond to hormone stimulation and production of viable oocytes. Notably, AMH is an indirect marker of the quiescent pool of primordial follicles that make up the ovarian reserve and is produced by the proliferating granulosa cells surrounding an oocyte during development, prior to ovulation. AMH has previously been measured in cohorts of endometriosis patients, most commonly those with ovarian endometrioma. In these cases, AMH often decreases after surgical removal of an endometrioma via cystectomy (Wang, Liu et al. 2020, Muraoka, Osuka et al. 2021, Sarbazi, Akbari et al. 2021, Fakehi, Davari Tanha et al. 2022, Mansouri, Safinataj et al. 2022, Shi, An et al. 2022, Tang and Li 2022, Crestani, Merlot et al. 2023). Some reports suggest AMH levels increase again during follow-up periods, however they never return to pre-surgery levels (Kostrzewa, Wilczyński et al. 2019, Sadullayev and Medvediev 2022). To date, only one study has reported AMH levels in a cohort of women with SPE, demonstrating no change in AMH compared to age-matched, population controls (n=62 in each group) (Lessans, Gilan et al. 2023). In another study, 40 expectantly managed women (no surgical intervention to treat their ovarian endometriosis) were reported to have AMH levels which declined faster than control women without endometriosis (Kasapoglu, Ata et al. 2018) providing the strongest evidence to date that there is a mechanism by which endometriosis drives infertility. In the current study, serum AMH was measured in a cohort of women with SPE, and levels were compared to both women without endometriosis (confirmed absence of lesions by laparoscopy), and a group of women also with an endometrioma (SPE+OE), to investigate how AMH may be altered by SPE. Additionally, we measured systemic and peritoneal inflammation as potential mechanisms driving endometriosis-associated infertility and associated changes to AMH levels.

## Materials and Methods

### Ethical approval

Participants were recruited in South-East Scotland between October 2015 and July 2023 from the Royal Infirmary of Edinburgh (NHS Lothian, REC 20/LO/1298) and gave informed consent to be involved in the study and for biospecimens to be collected during surgery.

### Participants samples

Participants were scheduled to undergo diagnostic laparoscopy for suspected endometriosis, or other gynaecological procedures (‘no endo’ group only). A venous blood sample was taken prior to surgery, and where possible, a peritoneal fluid sample collected during surgery.

Biospecimens were processed in accordance with WERF EPHect protocols (Rahmioglu, Fassbender et al. 2014). Serum was collected from whole blood samples via centrifugation at 2500g for 10 minutes at 4°C and aliquots stored at −80°C. Peritoneal fluid was similarly processed by centrifugation at 900g for 5 minutes at 4°C and aliquots stored at −80°C.

### Clinical information

Participants were categorised by an endometriosis surgeon according to surgical findings; no endometriosis (no lesions observed; no endo), superficial peritoneal endometriosis (only superficial lesions present; SPE), or superficial peritoneal endometriosis with an ovarian endometrioma (SPE+OE) (Table 1). Exclusion criteria included participants currently pregnant or breastfeeding, known reproductive malignancies, and previous history of endometriosis, specifically endometrioma. Current use of hormonal treatments to manage endometriosis symptoms was used to stratify samples. Hormone treatments included the combined oral contraceptive pill (COCP), progesterone only pill (POP), Depo-Provera^TM^, levonorgestrel intrauterine system (LNG-IUS), Nexplanon^TM^ implant, norethisterone, or a contraceptive patch. Their prevalence of use in this cohort is summarised in Table 1. The Endometriosis Fertility Index (EFI) was calculated for each participant as previously described (Adamson and Pasta 2010).

**Table 1.** Participant hormone treatment use.

|  | **No endo**<br>**% (n)** | **SPE**<br>**% (n)** | **SPE+OMA**<br>**% (n)** |
| --- | --- | --- | --- |
| *COCP* | 10.25 (4) | 11.63 (5) | 4.34 (1) |
| *POP* | 20.50 (8) | 16.27 (7) | 4.34 (1) |
| *LNGIUS* | 12.82 (5) | 18.60 (8) | 13.04 (3) |
| *Depo-Provera* | 10.25 (4) | 0.00 (0) | 4.34 (1) |
| *Other* | 10.25 (4) | 6.97 (3) | 0.00 (0) |
| *No hormones* | 35.89 (14) | 46.51 (20) | 73.91 (17) |

No endo – no endometriosis lesions visualised at laparoscopy, SPE – superficial peritoneal endometriosis, SPE+OMA – superficial peritoneal and ovarian endometriosis, COCP – combined oral contraceptive pill, POP – progesterone only pill, LNGIUS – levonorgestrel intrauterine system, other includes norethisterone, patch, Nexplanon implant.

### AMH ELISA

Serum samples were diluted 1:10 and assayed in duplicate using the human picoAMH ELISA (AL-124-r, Ansh Labs, United States) according to manufacturer’s directions. Absorbance was read using Clariostar plate reader (BMG Labtech) at 450nm.

### Luminex

Circulating inflammatory cytokines were quantified using custom 13-plex Discovery Luminex (Bio-Techne, United States) to detect CD163, IL-1β, IL-8/CXCL8, IL-23, MIF, TGF-α, β-NGF, IL-1α, IL-6, IL-17A, IL-33, NRG1, and TNF-α according to manufacturer’s instructions. Serum samples were diluted 1:2, and peritoneal fluid diluted 1:10. All samples were assayed in duplicate. Luminex detection was completed using a Luminex xMAP INTELLIFLEX analyser using LX200 low sensitivity setting.

### PGE2 ELISA

For measurement of prostaglandin E2 levels serum and peritoneal fluid were diluted 1:2 and assayed in duplicate according to manufacturer’s instructions (500141, Cayman Chemical, United States). Absorbance was read using Clariostar plate reader (BMG Labtech) at 420nm.

### CD14 ELISA

Serum and peritoneal fluid samples were diluted 1:2 and assayed in duplicated according to manufacturer’s instruction (A75809, antibodies.com, United Kingdom). Absorbance was read using Clariostar plate reader (BMG Labtech) at 450nm.

### CCL18 ELISA

Serum and peritoneal fluid samples were diluted 1:100 and assayed in duplicate according to manufacturer’s instruction (DCL180B, RCD Systems, United States). Absorbance was read using Clariostar plate reader (BMG Labtech) at 450nm.

### Statistical analysis

Analyses were completed in GraphPad Prism 10. ELISA results were interpolated from standard curve generated from each assay’s unique standards and corresponding blank corrected absorbance values. Luminex analyses were completed using Quantist software (BioTechne, United States). Data were assessed for normality using Shapiro-Wilk normality test and the appropriate statistical test chosen accordingly. Linear regression modelling was performed using RStudio (Version 2024.12.1+563) using lm() function within the ‘stats’ package including a two-way interaction term to assess the relationship between endometriosis subtypes and hormone use, with age as a covariate. Comparisons of two groups used Welch’s t-test for normal distribution, or Mann Whitney U test for non-normally distributed data. Comparison of three groups utilised one-way ANOVA with Tukey’s post-hoc test for normal distribution, or Kruskal-Wallis with Dunn’s post-hoc test for non-normal distribution.

## Results

### Women with superficial endometriosis have lower EFI and serum AMH levels compared to women without endometriosis

Demographic information including age and BMI of participants and their use of hormone treatments are summarised in Tables 1 and 2. There was no difference in age between the no endometriosis and SPE groups, but the SPE+OE women were older than both no endometriosis and SPE women (Table 2, Figure 1A). Body mass index (BMI) was comparable between all groups (Table 2, Figure 1B). Most women had no history of being diagnosed with infertility and 43-60% of women in each group had been pregnant in the past (Table 2). Women with SPE had lower EFI scores than women with no endometriosis (Table 3, Figure 1C), while SPE+OE women had lower EFI scores compared to both women with no endometriosis, and those with SPE alone (Figure 1C). Serum AMH levels for SPE+OE were lower than no endometriosis women (Table 3), however linear regression modelling was performed to allow AMH levels between groups to be compared while accounting for confounding variables like age and use of hormone treatments (Table 4). Both SPE and SPE+OE groups had significantly lower AMH levels compared to the no endometriosis control when controlling for participant age (p=0.009 and p=0.002, respectively; Table 4). When accounting for both age and hormone use, SPE+OE women had significantly lower AMH levels (p=0.02), while women with SPE alone did not (p=0.069), suggesting hormone use may contribute to differences observed in AMH.

**Figure 1.**
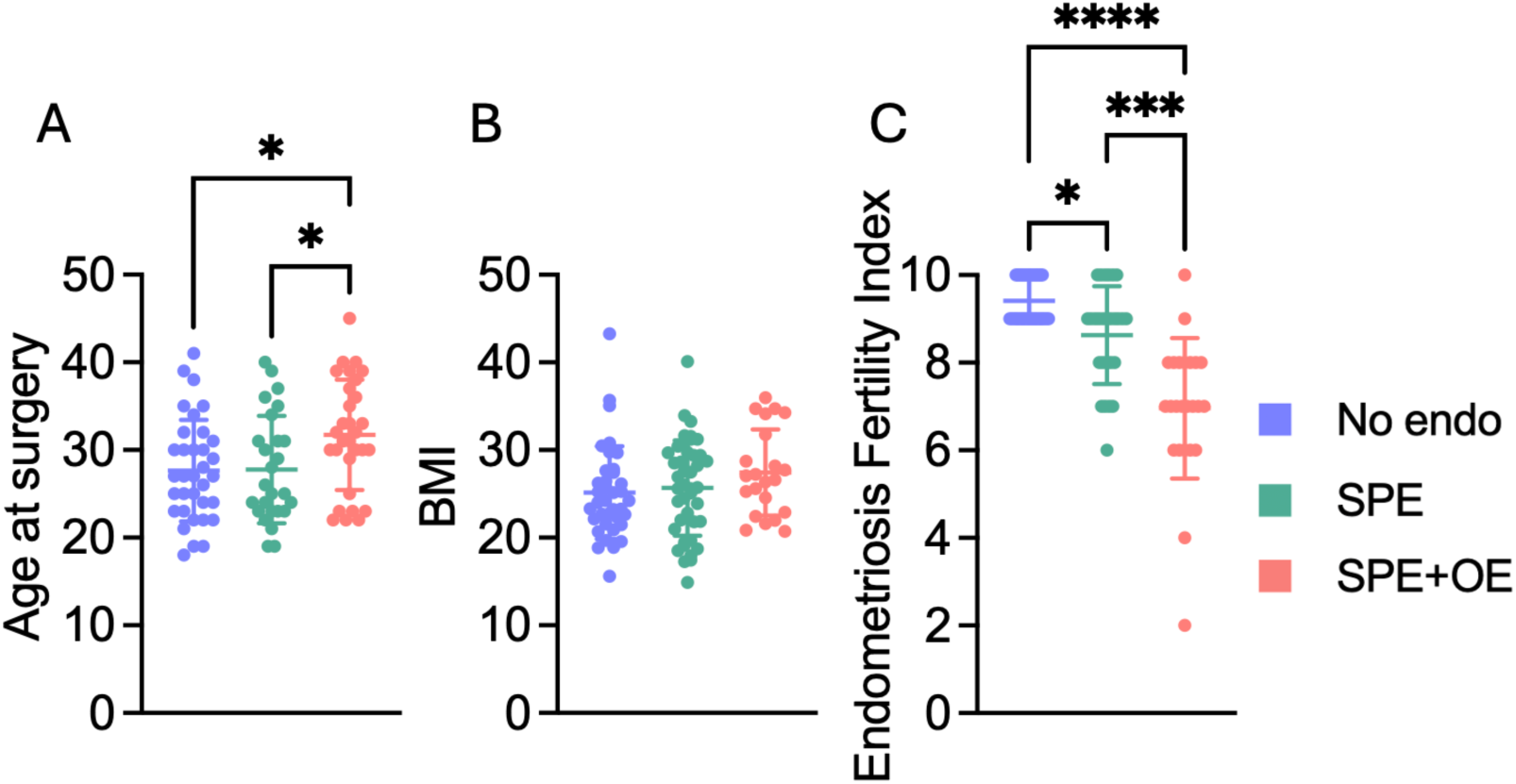
Women with SPE have impaired fertility. A) SPE+OE women are older than no endo controls and those with SPE only (n=26-36), with BMI similar across groups (B; n=22-43). Endometriosis Fertility Index (EFI) is reduced for both SPE alone and SPE+OMA groups (C; n=24-29). Data are mean±SD, *p<0.05, **p<0.01, ***p<0.001, ****p<0.0001. Shapiro-Wilk test for normality, one-way ANOVA with Tukey’s post-hoc test (A) or Kruskal-Wallis with Dunn’s post-hoc test (B, C,).

**Table 2.** Participant demographic information.

|  | **No endo** | **SPE** | **SPE+OMA** |
| --- | --- | --- | --- |
| **Sample size (n)** |  |  |  |
| *Whole cohort* | 39 | 43 | 23 |
| *No hormones* | 14 | 20 | 17 |
| *Hormones* | 25 | 23 | 6 |
| **Age Mean±SD** | 27.65 ± 5.81 | 27.77 ± 6.14 | 31.73 ± 6.31*^ |
| **BMI Mean±SD** | 25.14 ± 5.29 | 25.68 ± 5.44 | 27.43 ± 4.92 |
| **History of infertility (n)** |  |  |  |
| *Yes* | 4 | 2 | 8 |
| *No* | 32 | 36 | 11 |
| *Information unavailable* | 3 | 5 | 4 |
| **History of pregnancy (n)** |  |  |  |
| *Yes* | 16 | 26 | 10 |
| *No* | 19 | 14 | 10 |
| *Information unavailable* | 4 | 3 | 3 |

No endo – no endometriosis lesions visualised at laparoscopy, SPE – superficial peritoneal endometriosis, SPE+OMA – superficial peritoneal and ovarian endometriosis, BMI – body mass index, SD – standard deviation. \*p<0.05 vs no endo, \*\*p<0.01 vs no endo, \*\*\*p<0.001 vs no endo, ^p<0.05 vs SPE.

**Table 3.** Endometriosis fertility index and levels of circulating AMH. Mean*±*SD or median [IQR].

|  | No endo | SPE | SPE+OMA |
| --- | --- | --- | --- |
| **EFI Mean±SD** | 9.41 ± 0.50 (29) | 8.63 ± 1.11 (27) | 6.95 ± 1.60 (24) |
| **AMH (ng/mL)** |  |  |  |
| *Median [IQR] (n)* | 1.15 [0.75, 1.94] (36) | 0.89 [0.53, 1.52] (41) | 0.73 [0.32, 1.19] (29) |
| *Mean ± SD (n)* | 1.29 ± 0.80 (36) | 1.02 ± 0.66 (41) | 0.77 ± 0.52 (29) |

No endo – no endometriosis lesions visualised at laparoscopy, SPE – superficial peritoneal endometriosis, SPE+OMA – superficial peritoneal and ovarian endometriosis, EFI – endometriosis fertility index, AMH – anti-Mullerian hormone, SD – standard deviation, IQR – interquartile range.

**Table 4.** Linear regression model of AMH. Relationship between AMH and SPE/SPE+OMA vs control, with age or age and hormone use taken into consideration. *p<0.05, **p<0.01.

| Predictor | Estimate | Standard error | t-value | p-value | Significance |
| --- | --- | --- | --- | --- | --- |
| SPE+age | -0.64 | 0.24 | -2.68 | 0.009 | ** |
| SPE+OMA+age | -0.77 | 0.25 | -3.12 | 0.002 | ** |
| SPE+age+hormones | 0.57 | 0.31 | 1.83 | 0.069 | ns |
| SPE+OMA+age+hormones | 0.96 | 0.43 | 2.21 | 0.02 | * |

### Elevated inffammatory cytokines observed in circulation of women with SPE

We next sought to understand potential driving mechanisms of this change in AMH with SPE and turned to inflammation as a hallmark of endometriosis. Inflammatory cytokines and proteins were quantified using multiplex Luminex and ELISA’s on the same serum samples used to measure AMH and are summarised in Table 5. When cohorts were separated by use of hormones for the majority of cytokines in serum were not significantly changed between SPE and no endometriosis groups (Table 5). The two exceptions were IL-17 (Figure 2A) and TNF-α (Figure 2B) both of which were significantly increased in women with SPE not using hormones (n=9) compared to controls (n=6); notably these differences were not significant is women were using hormone therapies.

**Figure 2.**
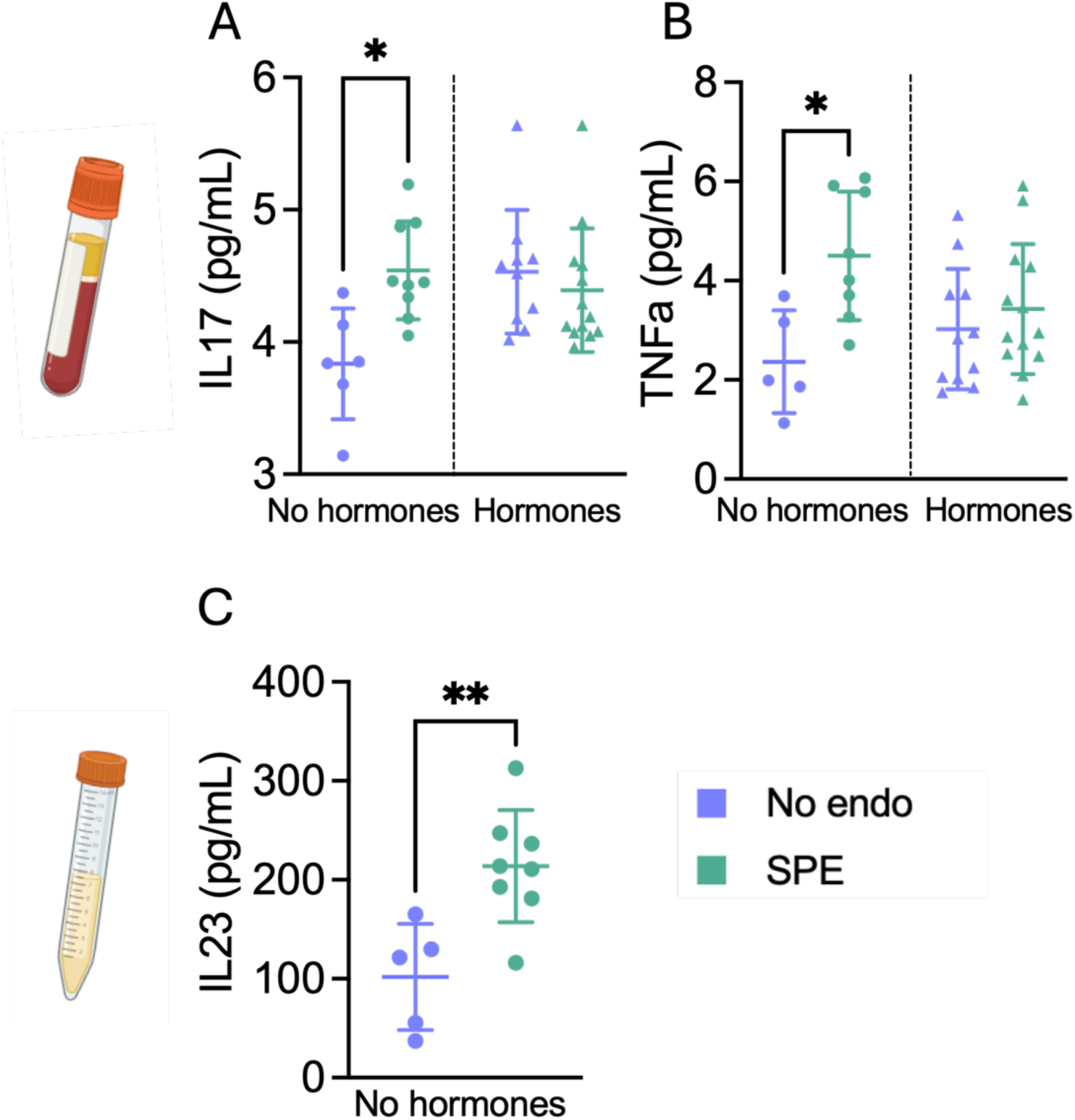
Women with SPE have elevated inflammatory cytokines, seen only in women not using hormone treatments. Levels of IL17 (A) and TNFa (B) in the circulation, and IL23 (C) in the peritoneal fluid are elevated in women with SPE not using hormones, compared to no endo. Data are mean±SD, n=6-13/group, *p<0.05, **p<0.01. Shapiro-Wilk test for normality, one-way ANOVA with Tukey’s post-hoc test (A, B), or Welch’s t-test (C) for significance.

**Table 5.** Cytokine levels in the circulation of women with and without superficial endometriosis.

| **Cytokine** | **Hormone treatment** | **No endo (n=17)** | **SPE (n=22)** |
| --- | --- | --- | --- |
| βNGF (pg/mL) | No hormones | 2.67 [2.25, 3.92] | 3.87 [3.18, 4.50] |
|  | Hormones | 2.96 [2.82, 4.42] | 3.70 [2.47, 4.16] |
| CCL18 (ng/mL) | No hormones | 588.1 [324.1, 773.9] | 641.6 [390.0, 770.3] |
|  | Hormones | 486.9 [367.6, 557.1] | 363.8 [307.0, 570.5] |
| CD14 (ng/mL) | No hormones | 4.07 [3.44, 5.69] | 4.14 [3.10, 5.36] |
|  | Hormones | 3.19 [2.68, 3.45] | 3.60 [2.96, 4.96] |
| CD163 (ng/mL) | No hormones | 804.8 [196.7, 942.4] | 358.3 [305.4, 963.3] |
|  | Hormones | 447.8 [208.3, 566.4] | 613.1 [313.9, 808.6] |
| IL-1α (pg/mL) | No hormones | 0.0 [0.0, 0.03] | 0.0 [0.0, 0.18] |
|  | Hormones | 0.0 [0.0, 0.0] | 0.0 [0.0, 0.12] |
| IL-1β (pg/mL) | No hormones | Undetected | Undetected |
|  | Hormones | Undetected | Undetected |
| IL-6 (pg/mL) | No hormones | 0.49 [0.07, 1.73] | 0.54 [0.07, 4.16] |
|  | Hormones | 0.56 [0.13, 1.06] | 0.22 [0.09, 1.66] |
| IL-8 (pg/mL) | No hormones | 8.97 [4.60, 29.59] | 7.34 [3.53, 13.51] |
|  | Hormones | 5.00 [3.55, 6.77] | 6.30 [1.68, 12.74] |
| IL-17 (pg/mL) | No hormones | 3.84 [3.54, 4.19] | 4.45 [4.26, 4.88]* |
|  | Hormones | 4.55 [4.15, 4.66] | 4.19 [4.07, 4.59] |
| IL-23 (pg/mL) | No hormones | Undetected | 0.0 [0.0, 15.63] |
|  | Hormones | 0.0 [0.0, 32.8] | 0.0 [0.0, 17.7] |
| IL-33 (pg/mL) | No hormones | 0.67 [0.22, 7.85] | 1.23 [0.52, 1.73] |
|  | Hormones | 0.93 [0.06, 9.40] | 0.89 [0.18, 25.14] |
| MIF (pg/mL) | No hormones | 662.9 [0.0, 1561] | 0.0 [0.0, 3713] |
|  | Hormones | 1029 [341.5, 1341] | 506.8 [138.9, 1355] |
| NRG1 (pg/mL) | No hormones | 0.0 [0.0, 141.8] | Undetected |
|  | Hormones | 1.58 [0.0, 22.61] | 0.0 [0.0, 22.77] |
| PGE2 (ng/mL) | No hormones | 337.3 [187.0, 654.0] | 215.1 [74.46, 403.4] |
|  | Hormones | 893.4 [469.6, 2203] | 378.1 [178.6, 707.8] |
| TGF-α (pg/mL) | No hormones | 7.79 [3.48, 17.93] | 9.58 [6.42, 16.11] |
|  | Hormones | 9.99 [8.39, 18.49] | 13.88 [9.91, 16.83] |
| TNF-α (pg/mL) | No hormones | 1.99 [1.49, 3.43] | 4.28 [3.37, 5.88]* |
|  | Hormones | 2.83 [2.03, 3.73] | 2.95 [2.50, 4.35] |

No endo – no endometriosis lesions visualised at laparoscopy, SPE – superficial peritoneal endometriosis, IQR – interquartile range. Median [IQR], \*p<0.05 vs no endo.

### Inffammation in the local peritoneal microenvironment differs from the circulation

In a subset of women (n=15), pelvic peritoneal fluid samples from those not using hormones were used to assess inflammation levels in the local pelvic peritoneal microenvironment. Concentrations of peritoneal fluid inflammatory markers are summarised in Table 6. Whilst most factors were detected, variation in levels meant most did not reach statistical significance, including those previously shown to be elevated in serum. Interestingly, one exception was IL-23 that was significantly elevated in women with SPE not using hormones compared to those without endometriosis (Figure 2C).

**Table 6.**
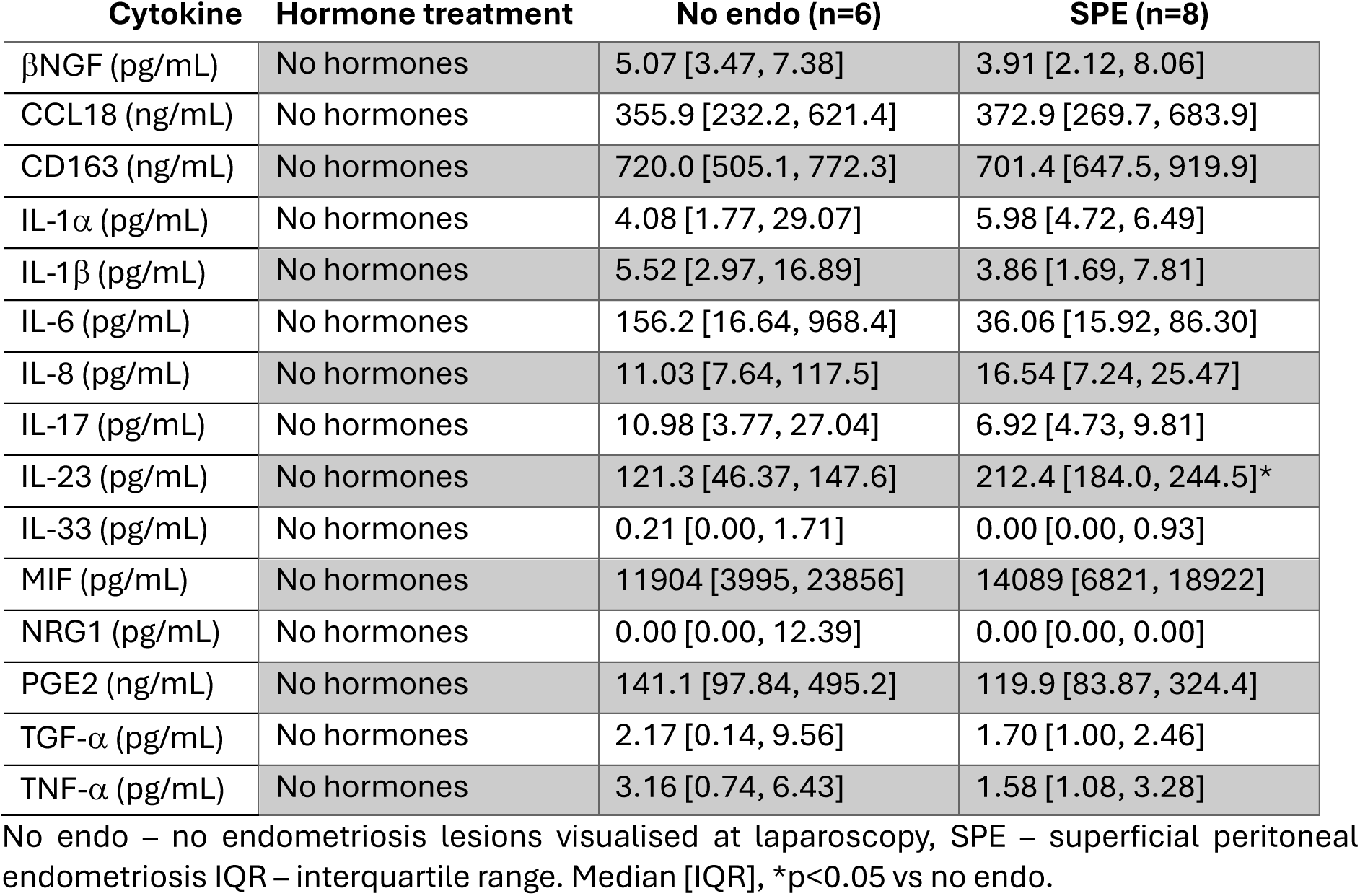
Cytokine levels in the peritoneal fluid from women with and without superficial endometriosis.

## Discussion

The mechanisms underlying endometriosis-associated infertility are poorly understood and rarely characterised by subtype of endometriosis. Here, we demonstrate for the first time that serum AMH is decreased in women who only have lesions identified as the SPE subtype, compared to women confirmed to have no endometriosis lesions detected during surgery. Interestingly, this finding was associated with elevated pro-inflammatory cytokine levels in the same biospecimens, suggesting inflammation is one mechanism contributing to endometriosis-associated delays in conception in women with SPE.

Notably, in our cohort, we found the EFI was lower for women with SPE or SPE+OE compared to no endometriosis controls, suggesting a lower probability of future pregnancy success for women with either subtype of endometriosis even though some had already achieved a pregnancy. Since its inception, the EFI has been validated extensively and can be reliably and routinely calculated to predict pregnancy success for non-ART conceptions (Adamson and Pasta 2010, Tomassetti, Geysenbergh et al. 2013, Vesali, Razavi et al. 2020). However, until our study, it had not been used specifically to evaluate future fertility in women with endometriosis by making a direct comparison to a similar cohort of women without endometriosis. The fact the EFI was reduced in our cohort of women with SPE is supported by a recent population study from Finland demonstrating women with endometriosis have a lower overall fertility rate compared to women without endometriosis (Tuominen, Saavalainen et al. 2025). Notably, in their data was a historical population-based cohort study (1998-2012) with data from 18,320 fertile-aged women with a surgical diagnosis of endometriosis; 5786 of whom had a diagnosis of SPE alone. They reported lower fertility rates in the endometriosis group compared to a reference group. Although, consistent with our data, 68% did achieve a pregnancy during follow up. While registry studies provide large datasets, they are not able to provide insight on whether their lower fertility rate is due to an underlying biological reason or less sexual activity due to debilitating symptoms associated with endometriosis. Our study starts to bridge this gap by providing evidence of lower EFI and serum AMH in the same women with SPE alone, or SPE with OE compared to no endometriosis controls.

In this study, we focused on SPE as this is the most common subtype of endometriosis and one that receives less attention in fertility studies than ovarian disease. We also focus on the impact to ovarian function and fertility, rather than the endometrium as this has already been the subject of several previous studies (reviewed (Griffiths, Horne et al. 2024). A strength of our cohort is the precise phenotyping of the participants and their disease subtypes. Previous studies have assessed fertility in groups of women with endometriosis, irrespective of the subtype (reviewed (Griffiths, Horne et al. 2024). Our cohort was stratified into groups based on their surgical findings and medical history to confirm the absence of any previous diagnosis of an endometrioma that could confound the AMH results. Moreover, the control ‘no endo’ population has advantages over previously reported studies as women in this group were confirmed as having no observable endometriosis lesions after undergoing an identical diagnostic laparoscopy to those confirmed to have endometriosis lesions. In contrast, previous studies investigating endometriosis-associated infertility have either utilised age-matched controls where the presence of asymptomatic and undiagnosed endometriosis has not been ruled out surgically, or women attending fertility clinics (Lessans, Gilan et al. 2023). In both cases there are likely to be other factors contributing to infertility, and as such neither is an ideal control (‘fertile’) group to compare to the fertility in a group of women with endometriosis. This concern is also based on reports describing that almost half of women presenting to fertility clinics with unexplained infertility are subsequently found to have endometriosis, most of which have SPE (Van Gestel, Bafort et al. 2024).

Interestingly, changes in AMH levels were influenced by hormone use and a significant reduction in AMH levels for women with SPE was not seen when hormone use was taken into account in linear regression modelling. The ability of hormone treatments to alter AMH has been reported previously (Bernardi, Weiss et al. 2021, Hariton, Shirazi et al. 2021, Nelson, Ewing et al. 2023), however, the influence of hormone use on AMH levels for women who also have endometriosis has never been investigated to the best of our knowledge. It is possible the lack of change here is also driven by a smaller sample size once participants are stratified by their use of hormones. Therefore, studies in a larger sample size are warranted to confirm and validate these findings.

To investigate a potential mechanism by which the pelvic microenvironment may contribute to depleted AMH, the associated inflammatory environment in the peritoneal fluid recovered at the time of surgery from the same women was characterised. From the panel of cytokines investigated here, which were selected based on previous works and evidence in the literature, only IL-17 and TNF-α, and IL-23 were found to be elevated with SPE in the serum and peritoneal fluid, respectively. These results align with previous works demonstrating a dysregulated IL-17/IL-23 axis, and elevated TNF-α levels with endometriosis (Harada, Yoshioka et al. 1997, Sisnett, Zutautas et al. 2024). For example, Sisnett et al. (2024) showed elevated IL-23 in the plasma of patients with endometriosis (n=13) compared to healthy, fertile controls (n=19) (Sisnett, Zutautas et al. 2024). Elevated levels of TNF-α (Harada, Yoshioka et al. 1997) and IL-17 (Zhang, Xu et al. 2005) in the peritoneal fluid of women with endometriosis has been previously reported. A strength of our study over previous work is our subgroup analysis focussed on SPE or SPE+OMA, rather than a general endometriosis versus control comparison. Interestingly, Zhang et al. (2005) also reported an even greater increase in IL-17 levels for patients with endometriosis and a diagnosis of infertility compared to endometriosis alone (Zhang, Xu et al. 2005). Due to limited sample size, we were unable to complete the same analyses. Taken together with our study showing IL-17 is elevated in the same biospecimens where AMH and EFI are reduced, it would be interesting for future studies to attempt to disentangle the relationship between IL-17 and endometriosis pathogenesis and endometriosis-associated infertility.

Interestingly, differences in circulating IL-17 and TNF-α were unique to the participants not using hormone treatments. Interestingly, this appeared to be due to a similar elevation in these cytokines in the no endometriosis group using hormone treatments. While enhanced inflammatory responses to acute stressors have been reported previously in women using hormone treatments (Larsen, Cox et al. 2020, Mengelkoch, Gassen et al. 2024), to the best of our knowledge this has not been assessed in women with a more chronic inflammatory profile, as with endometriosis. Moreover, it is unclear what the consequences may be on levels of proinflammatory cytokines and subsequently AMH levels for women with SPE or SPE+OE if they stop taking hormone treatments to attempt to conceive. These are all relevant avenues for future work characterising how inflammation contributes to endometriosis-associated infertility.

Surprisingly, in our study no differences were observed in well-established factors known to play a role in endometriosis-associated symptoms including PGE2 (Rakhila, Carli et al. 2013) and IL-8 (Jørgensen, Hill et al. 2017). This may be due to differences in the biospecimens investigated with the Rakhila (2013) study focused on levels in ectopic lesion tissue. The study from Jørgensen and colleagues (2017) measured levels of peritoneal fluid and reported significantly higher levels of IL-8 in 56 endometriosis patients compared to 38 without endometriosis attending a fertility clinic in biospecimens recovered during the luteal/secretory phase. In our study, we had only 8 samples from women not on hormones and the increase detected compared with controls was not statistically significant.

IL-33 has been studied in endometriosis previously and shown to be elevated with deep endometriosis (Santulli, Borghese et al. 2012, Mbarik, Kaabachi et al. 2015, Miller, Monsanto et al. 2017). This may explain why it is unchanged in our study, as patients with deep endometriosis were not investigated. Mbarik et al. (2015) divided their cohort by the revised American Society for Reproductive Medicine staging and found stage I-II endometriosis (closest equivalent to the SPE group in the current study) had similar circulating IL-33 levels compared to control but elevated levels for the stage III-IV group in both the serum and peritoneal fluid (Mbarik, Kaabachi et al. 2015). A limitation to our study is also one of its aforementioned strengths. While our control group are confirmed to have no endometriosis lesions visualised at the time of laparoscopy, they all experience pelvic pain justifying their investigation for suspected endometriosis. It is possible alterations in the levels of these cytokines may be driven by inflammatory processes similarly contributing to pelvic pain which may explain the lack of differences observed between groups here, particularly for the cytokines previously extensively published in relation to endometriosis. It is also possible these inflammatory peritoneal processes present for those with chronic pelvic pain contribute to infertility. Serum AMH levels in our ‘no endo’ control group were lower than those reported in previous literature. The mean value for our ‘no endometriosis’ control group was 1.15ng/mL (compared to 0.89ng/mL in the SPE group). The previous study by Lessans et al. (2023) reported a mean AMH value of 3.0ng/mL for their age-matched control group (2.8ng/mL for peritoneal endometriosis), and a study combining all subtypes of endometriosis reported mean AMH of 2.30ng/mL for healthy controls (1.99ng/mL for endometriosis) (Lessans, Gilan et al. 2023, Ramezani Tehrani, Mousavi et al. 2025). Together, these data may suggest a mechanism where the pelvic microenvironment (including pelvic inflammation), irrespective of presence of endometriosis lesions contributes to diminished AMH and subfertility. Women with other pelvic inflammatory conditions such as Crohn’s disease or inflammatory bowel disease are known to impact fertility if they have a flare up of their condition during attempts to conceive and during pregnancy itself (Nguyen, Seow et al. 2016, Mahadevan, Robinson et al. 2019, Rosiou and Selinger 2023, Torres, Chaparro et al. 2023). While this requires further investigation in the settings of pelvic pain and endometriosis, the notion of an inflammatory peritoneum compromising fertility is plausible.

## Conclusion

Here, for the first time, we demonstrate EFI scores and AMH levels are reduced for women with SPE compared to women without endometriosis. These changes are accompanied by greater levels of IL-17 and TNF-α in the serum, and IL-23 in the peritoneal fluid. Interestingly, these differences were only observed in women not using hormone treatments. Our findings provide valuable information for clinicians to utilise when counselling women with endometriosis about their future fertility, with or without an endometrioma present.

## Data Availability

All non-identified data in the present study can be shared upon reasonable request to the authors.

## Acknowledgements

The authors would like to thank the EXPPECT Edinburgh clinical team for study recruitment and sample collection, and specifically Dr Lucy Whitaker for clinical input. Also, thanks to the expertise from the University of Edinburgh SURF Biomolecular & Assay Core, specifically Dr Kirsten Wilson and Linda Ferguson for Luminex technical assistance.

## Author’s roles

Study design: MJG, DAG, PTKS, AWH. Experimental procedures, statistical analysis: MJG, MB, FC. Results interpretation: MJG, DAG, PTKS, CED, AWH. Manuscript writing: MJG, DAG, PTKS, AWH. Manuscript editing: MJG, MB, FC, DAG, CED, PTKS, AWH.

